# TCMM: A Unified Database for Traditional Chinese Medicine Modernization and Therapeutic Innovations

**DOI:** 10.1101/2024.02.19.24303055

**Authors:** Zhixiang Ren, Yiming Ren, Zeting Li, Huan Xu

## Abstract

Mining the potential of traditional Chinese medicine (TCM) in treating modern diseases requires a profound understanding of its action mechanism and a comprehensive knowledge system that seamlessly bridges modern medical insights with traditional theories. However, existing databases for modernizing TCM are plagued by varying degrees of information loss, which impede the multidimensional dissection of pharmacological effects. To address this challenge, we introduce traditional Chinese medicine modernization (TCMM), the currently largest modernized TCM database that integrates pioneering intelligent pipelines. By aligning high-quality TCM and Western medicine data, TCMM boasts the most extensive TCM modernization knowledge, including 20 types of modernized TCM concepts such as prescription, ingredient, target and 46 biological relations among them, totaling 3,447,023 records. We demonstrate the efficacy and reliability of TCMM with two features, prescription generation and knowledge discovery, the outcomes show consistency with biological experimental results. A publicly available web interface is at https://www.tcmm.net.cn/.

## 1. Introduction

TCM is an empirical science based on thousands of years of clinical experience and continues to play a crucial role in disease diagnosis and treatment[1]. In recent years, a wealth of studies[2, 3, 4, 5] have demonstrated that TCM and Western medicine share the same theoretical foundation at the molecular level. Specifically, compounds can treat diseases by regulating the efficacy of targets. Despite this, current information on the chemical components, and metabolism mechanisms of TCM is still lacking, making the relation between drug components and pharmacological effects unclear, hindering the targeted TCM diagnosis and treatment. Therefore, in the modernization process of TCM, it is urgent to establish a comprehensive and highly reliable TCM modernization database, which would facilitate the identification of active ingredient within herb and elucidate the mechanism of action with biological targets.

Recently, numerous efforts[6, 7, 8, 9] have been made to construct TCM databases that incorporate information about herb, ingredient, and target to support related research. However, these works often overlook the critical concepts of “symptom” and “prescription” inherent in the theoretical system of TCM. SymMap[10] incorporates symptom information, which is beneficial for phenotype-based drug discovery research. LTM-TCM[11] and CPMCP[12] introduce the concept of prescription, which helps Chinese medicine practitioners understand the compatibility of herbs in prescriptions. TCMBANK[13] integrates a vast amount of data on herb, ingredient, disease, and target entities but neglects important concepts like prescription and symptom in the theoretical system of TCM. Despite these advancements, current integrative works on TCM and Western medicine have only partially involved modern medical concepts like disease, target, and ingredient. This simplifies the principles of ingredient-based disease treatment and ignores essential information such as anatomy, and pathway, which is not conducive to understanding the complete pharmacological effects. Additionally, in the aforementioned databases, relations are binary, while in the biological process, relations between entities are complex and diverse. Different relations may represent the alleviation or aggravation of diseases, such as ingredient-downregulate/upregulate-gene.

Knowledge discovery is a hot topic in TCM, encompassing various challenging tasks such as digging into the molecular mechanisms of herbs and discovering the treatment patterns of TCM prescriptions. Although some researchers have used network pharmacology to study the action mechanism of prescriptions[14, 15, 16] and explore herb-symptom correlations[17], these efforts have focused on analyzing specific prescription or disease without discovering a universal method for explaining the mechanism. With advancements in artificial intelligence (AI), some methods combining TCM data and deep learning (DL) have been developed for tasks such as prescription-target interaction[18] and herb-symptom correlation[19]. However, due to the lack of detailed information, the pharmacological principle of TCM remains largely unexplored, leaving a gap in the knowledge discovery of modernized TCM.

Symptom-based prescription plays a crucial role in the treatment process of TCM, and the combination of prescription generation with AI methods has become a trend in recent years, providing convenience for patients and doctors. Recently, [20, 21] proposed the topic model for prescription generation that incorporates background knowledge such as herbal compatibility, aiming to facilitate the creation of prescriptions and integrate TCM theories into the generation process. To generate prescriptions that align more closely with TCM concepts, [22, 23, 24] focus on the use of seq2seq structures, by viewing TCM prescription generation as a sequence process task and decode the latent representation of symptoms into herb sequences. Additionally, GNN can effectively capture structural and semantic information between entities and is widely applied to prescription generation tasks. Many studies[25, 26, 27, 28, 29] have utilized GNN to capture high-order correlations among herbs, symptoms, and prescriptions, recommending highquality herb collections based on background knowledge. However, existing methods overlooked internal prescription information such as compatibility principles and inherent properties of medicinal materials, while also lacking the use of mechanistic information like pathways and targets. nn In this work, we introduce TCMM, a unified database for TCM modernization that integrates 6 high-quality databases of TCM and Western medicine. According to Table 1, TCMM is the currently largest non-commercial database for TCM modernization, consisting of 20 typical entities such as prescription, ingredient, target and 46 relations among them, aiming to cover the theoretical systems of TCM and Western medicine as comprehensively as possible. A web-based interface is provided for users to explore relations among herb, ingredient, target, and related pathway or disease. Based on the constructed database, a prescription generation pipeline with pre-train method is proposed, which creates highly credible prescriptions, thereby proving the high peformance of the database. This pipeline is integrated into the website to support user-customized prescription generation. Furthermore, in the absence of direct data, we pioneering use a multi-hop reasoning method to achieve TCM knowledge discovery. This includes 2 challenging tasks, prescription repositioning and symptom-related target prediction. The results highly match experimental conclusions in case studies, effectively supporting TCM modernization.

**Table 1.**
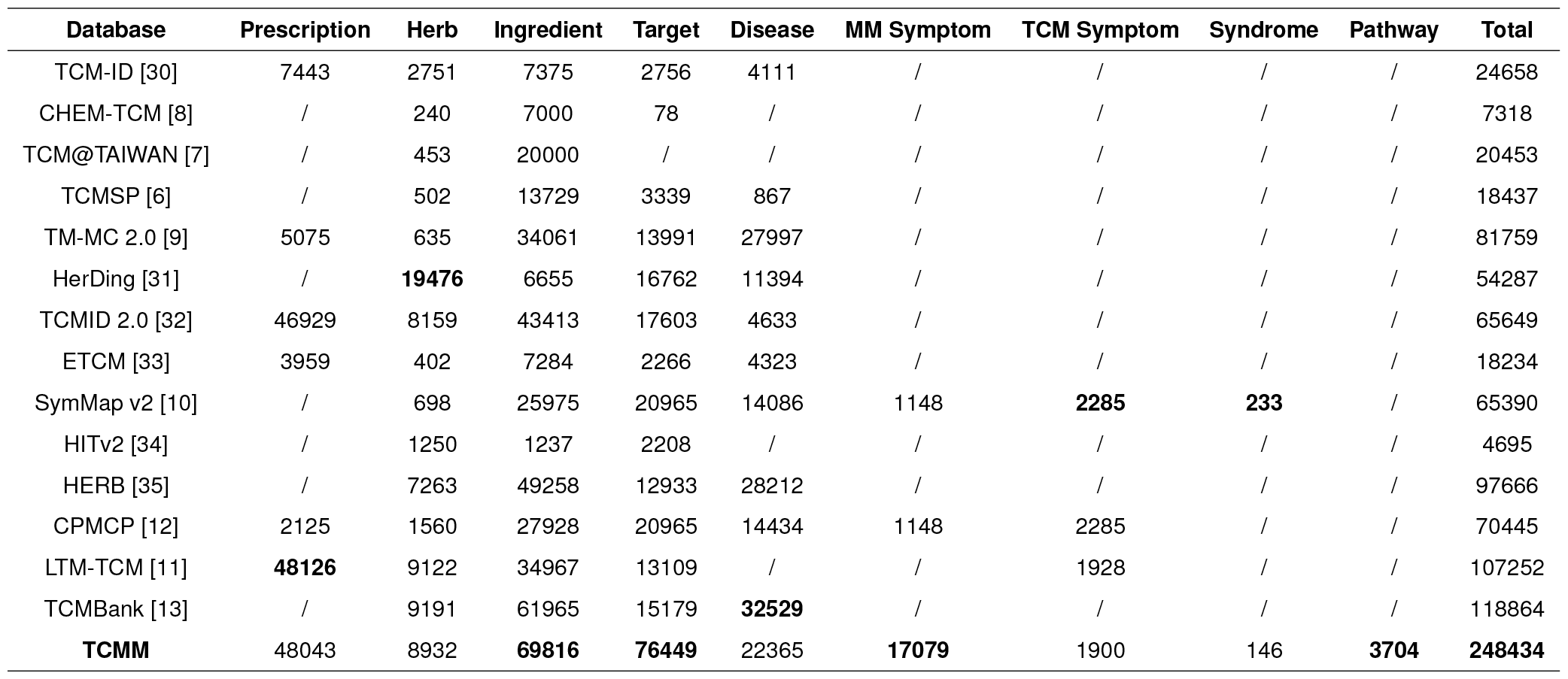
**TCM Database Comparison** provides a quantitative comparison of the data scale of TCMM with existing TCM databases. To ensure fairness, we only count the entity types present in existing databases. The values are based on the most recent data from the relevant databases. The results show that TCMM contains the most comprehensive entity type and the largest amount of data.

| Database | Prescription | Herb | Ingredient | Target | Disease | MM Symptom | TCM Symptom | Syndrome | Pathway | Total |
| --- | --- | --- | --- | --- | --- | --- | --- | --- | --- | --- |
| TCM-ID [30] | 7443 | 2751 | 7375 | 2756 | 4111 | / | / | / | / | 24658 |
| CHEM-TCM [8] | / | 240 | 7000 | 78 | / | / | / | / | / | 7318 |
| TCM@TAIWAN [7] | / | 453 | 20000 | / | / | / | / | / | / | 20453 |
| TCMSP [6] | / | 502 | 13729 | 3339 | 867 | / | / | / | / | 18437 |
| TM-MC 2.0 [9] | 5075 | 635 | 34061 | 13991 | 27997 | / | / | / | / | 81759 |
| HerDing [31] | / | **19476** | 6655 | 16762 | 11394 | / | / | / | / | 54287 |
| TCMID 2.0 [32] | 46929 | 8159 | 43413 | 17603 | 4633 | / | / | / | / | 65649 |
| ETCM [33] | 3959 | 402 | 7284 | 2266 | 4323 | / | / | / | / | 18234 |
| SymMap v2 [10] | / | 698 | 25975 | 20965 | 14086 | 1148 | **2285** | **233** | / | 65390 |
| HITv2 [34] | / | 1250 | 1237 | 2208 | / | / | / | / | / | 4695 |
| HERB [35] | / | 7263 | 49258 | 12933 | 28212 | / | / | / | / | 97666 |
| CPMCP [12] | 2125 | 1560 | 27928 | 20965 | 14434 | 1148 | 2285 | / | / | 70445 |
| LTM-TCM [11] | **48126** | 9122 | 34967 | 13109 | / | / | 1928 | / | / | 107252 |
| TCMBank [13] | / | 9191 | 61965 | 15179 | **32529** | / | / | / | / | 118864 |
| **TCMM** | **48043** | **8932** | **69816** | **76449** | **22365** | **17079** | **1900** | **146** | **3704** | **248434** |

## 2. Materials and methods

### 2.1. Collection and processing of TCMM

Existing TCM databases exhibit substantial differences in terms of processing methods and data sources, necessitating the establishment of independent alignment strategies for different types of entities and relationships. The processing methods for the main entities and relations within TCMM are described in detail as follows.

For entities, **Medicinal Material**, sourced from CPMCP and TCMBank, combines information from the “2015 Chinese Pharmacopoeia”, literature and multiple databases, such as TCMID, TCM-ID, SymMap, TCMSP, HERB, and TCM@TAIWAN. During data processing, records that share the same HERB ID or SymMap ID are merged. In instances where the attribute values differed, we combine the corresponding names separated by a semicolon. Subsequently, we extract attributes such as the “Four Qi and Five Flavors” and “Meridian Theory,” which encompassed 13 kinds of Tropism, 4 categories of Toxicity, 13 varieties of Flavor, and 12 types of Medicinal Properties.

**Prescription** comes from the TCMID and CPMCP, with the former derived from books and articles, and the latter’s from patent medicines and ancient prescriptions. We merge 2,140 prescriptions from CPMCP with 46,929 prescriptions from TCMID, resulting in 48,034 records after removing duplicates. Due to multiple names for some herbs in the prescriptions, we capture 4,947 alternative herb names from https://zhongyibaike.com and align the prescription information, in order to improve the recall rate of herb identification.

**Ingredient** comes from CPMCP and TCMBank. During the integration process, since CAS ID, PubChem ID, and InChI Key cannot uniquely determine the molecule, we merge data based on the same SymMap ID in TCMBank and CPMCP but remove records in TCMBank pointing to multiple SymMap IDs. For example, TCMBANKIN057994 points to SMIT00245 and SMIT1456. In this way, 69,816 ingredient records are ultimately obtained.

**Target** data is from CPMCP, TCMBANK, PharMeBINet, and PrimeKG databases. NCBI ID is used to integrate data from CPMCP, PrimeKG, and PharMeBINet, and supplement PrimeKG data with gene symbols. Furthermore, to better understand the role of targets in disease processes, we extract molecular function, biological process, and cellular component information for targets from PrimeKG and PharMeBINet and align them using GO ID.

**Symptom and syndrome** data comes from multiple databases. To explore the correlation between modern medicine (MM) and TCM diseases, MM symptoms and TCM symptoms are obtained separately. TCM symptoms come from CPMCP. Due to some overlapping semantics in TCM Symptoms and Syndromes in the source database, we utilize a large language model(LLM) to merge information from the source database and combine it with manual verification to improve accuracy. Specifically, the semantic descriptions of TCM Symptoms are first generated using prompt engineering combined with GPT methods[36, 37] and then convert the semantic descriptions into embedding representations using Sentence-BERT[38]. To reduce computation, symptoms are grouped according to locus and property and only calculate similarity within groups. Through manual verification, 0.98 is selected as the similarity threshold, and symptoms with a similarity greater than the threshold are merged into one record, resulting in 1,900 TCM Symptoms. To understand the cause of symptoms, TCMM also extracts Locus information from the CPMCP database as an attribute of TCM Symptoms. Syndrome information comes from the SymMap database, and 146 records are obtained after merging semantic information. In addition, 17,079 MM Symptom records are obtained by aligning information using shared MeSH IDs from PharMeBINet and CPMCP.

**Disease** comes from CPMCP, TCMBANK, PharMeBINet, and PrimeKG databases. We merge the data using keywords such as name, MONDO ID, OMIM ID, MeSH ID, Orphanet ID, and UMLS ID, resulting in 48,233 Disease records. Since a large number of phenotype information in TCMBANK, such as “Body Height” and “Cardiovascular Problem,” are categorized as diseases, we only retain Disease information from the PharMeBINet and PrimeKG databases in this study while preserving their database IDs in CPMCP and TCMBank.

For relations, **Prescription - Medicinal Material**: TCMM, by means of deep parsing of prescription descriptions and a combination of LLM with rule matching, extracts 321,913 relations between prescription and herbs from 48,034 prescriptions, of which 264,728 relations contained dosage information. Specifically, we employ ChatGLM to parse the medicinal materials and dosage information in the prescription descriptions. However, as the model is not trained with similar data, the accuracy is only 57%. To achieve better results, we manually annotate a dataset for fine-tuning ChatGLM and randomly extract 200 prediction results for manual evaluation, achieving an accuracy of 96% and a recall rate of 76%. For incorrectly predicted prescriptions, further predictions are made using rule matching, achieving an accuracy rate of 98% under the same evaluation method. In addition, we only retain 17 commonly used weight/volume units in herbal prescriptions and convert the ancient or imprecise units of herbal dosages to g or ml for the modernization of TCM. The dosage unit conversion method is shown in supplementary table S1.

**Prescription-Syndrome/Symptom**: The relation data of prescription-symptom partially originates directly from the CPMCP database, totaling 50,813 records, but this only covers part of the prescriptions. To further supplement information for other prescriptions, TCMM extracts symptom and syndrome information from the indication and treatment attributes of prescriptions in the TCMID and CPMCP databases. Specifically, we use the already obtained symptom and syndrome entities as keywords, match them with indication descriptions, and ultimately obtain an additional 60,342 prescription-symptom relations and 13,736 prescription-syndrome relations.

**Ingredient-Target**: Within TCMM, the Ingredient-Gene relation is refined into four different types: associate, bind, downregulate, and upregulate. The data for bind, downregulate and upregulate relations come from the PharMeBINet database, while the associate relation is obtained by integrating information from PharMeBINet, SymMap, CPMCP, and TCMBank databases. To better align with practical application scenarios, we establish a “mutual exclusion” rule for relations. Specifically, for the same ingredient-target group, downregulate and upregulate relations cannot coexist, and associates cannot coexist with the other three relations. For instance, in the PharMeBINet database, the same ingredient-target group was simultaneously marked as downregulate and upregulate. To resolve this conflict, we adopt the more broadly defined associate relation as a replacement.

**Target-Target**: TCMM contains three types of target-target relations, namely associate, regulate, and covary. The information for regulate and covary comes from the PharMeBINet database, while associate information comes from the PharMeBINet and PrimeKG databases. In TCMM, the associate relation is mutually exclusive with the other two relations, meaning that for the same target-target group, the associate will not coexist with either regulate or covary.

**Disease-Disease**: TCMM integrates resemble information from the PharMeBINet database. In this database, these two relations are deemed mutually exclusive. Therefore, for situations in the PharMeBINet database where two relations coexist, only the ‘is a’ information is retained.

**Ingredient-Disease**: The PharMeBINet database provides TCMM with induce, contraindicate, and treat relations for the ingredient-disease relation. In this database, since treat is semantically mutually exclusive with induce and contraindicate, we discard data from PharMeBINet where coexistence situations exist.

### 2.2. Data source and statistics of TCMM

TCMM is sourced from the 6 largest TCM knowledge bases, PrimeKG[39], PharMeBINet[40], TCMBank[13], SymMap[10], TCMID[32] and CPMCP[12], which results in a more detailed relations and a wider variety of entity types. In this work, the TCM or Western medical concepts, such as prescription, ingredient and target, are treated as entities. Additionally, a relation describes the biological correlation between entities. In this way, TCMM encompasses 20 types of entities and 46 kinds of relations, amounting to a total of 3,447,023 records, forming the most comprehensive TCM modernization database currently available. Data source of entities and relations is depicted in Figure 1, which integrates Western medicine information from PharMeBINet and PrimeKG, and integrates TCM information from TCMBank, CPMCP, and SymMap. Figure 2 presents detailed information and statistics of the database, which contains a total of 248434 entities and 3447023 triplets. The integration is beneficial for drug researchers to understand the pharmacological effects of TCM from a modern medical perspective. Additionally, it enhances the potential of the knowledge graph (KG), such as enabling it to address challenging tasks like prescription repositioning for modern diseases.

**Figure 1:**
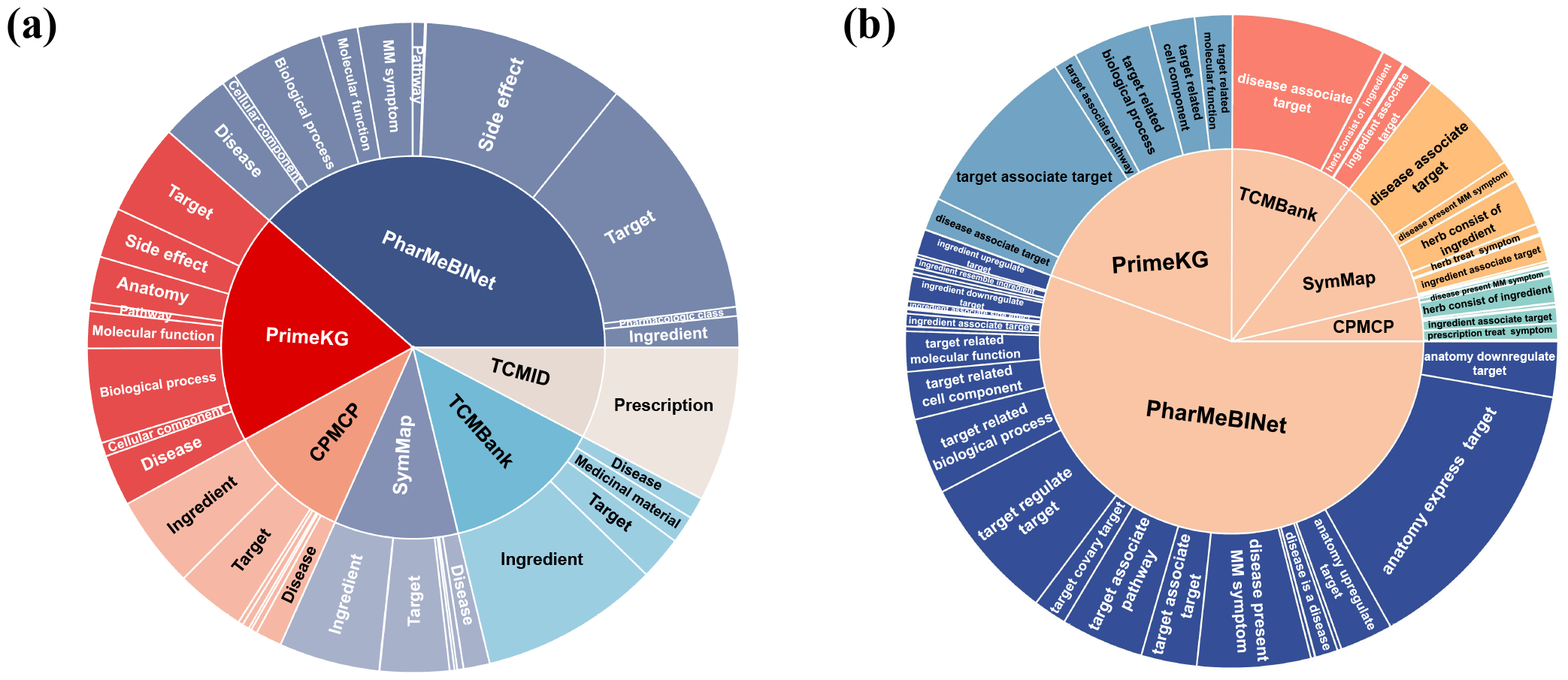
**Data Source of TCMM Database** provides a visual representation of the data sources of entities and relations in TCMM. The size of each area represents the quantity of entities or relations from that source. **(a), (b) Data Source of TCMM Entities and Relations** integrates Western medicine information from PharMeBINet, PrimeKG and integrates TCM information from TCMBank, CPMCP, TCMID and SymMap. Specific rules and LLM are used for information combination.

**Figure 2:**
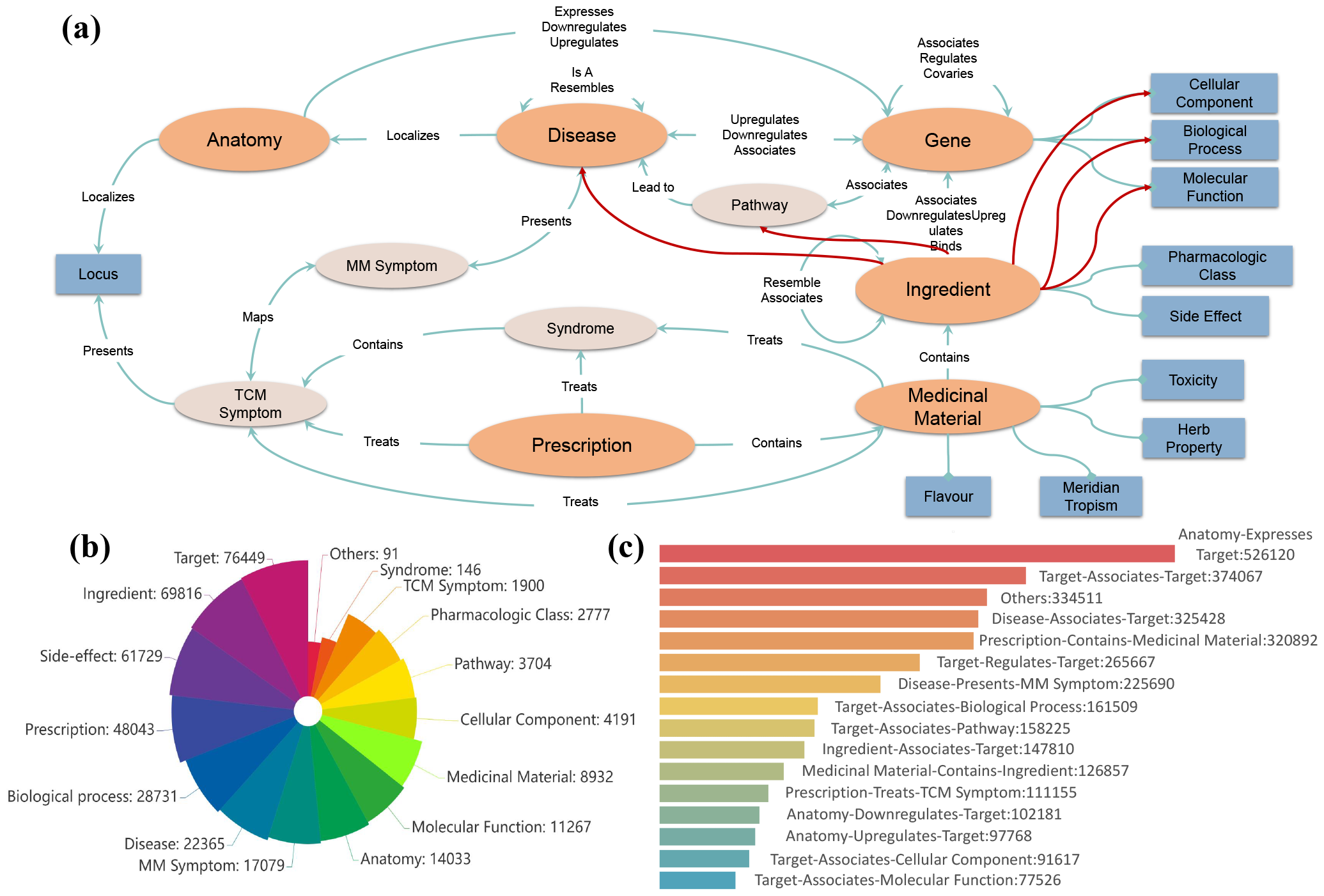
Overview of TCMM Database. **(a) Details of TCMM Database** displays the complete 20 entities and 46 relations in the database, while the blue bars represent the attribute entities. **(b) Entity Distribution in TCMM** provides a quantitative overview of the entity types in TCMM. TCM attribute entities are a small minority and are therefore grouped together as “others”. **(c) Relation Distribution in TCMM** illustrates the quantity of each relation type within the TCMM. 31 types of relations, due to their low frequency, have been collectively categorized as ‘others’

### 2.3. Knowledge augmentation based TCM prescription generation

In this study, we implement a TCM prescription generation pipeline upon the work proposed by [42],which grounds in the Encoder-Decoder framework shown in Figure 3. By combining it with the Diverse Beam Search[43], the model is proven to be capable of producing more diverse prescriptions. Furthermore, we extract relevant information from the database to construct a KG and incorporate Chinese and Western medical knowledge into the model via a graph pre-train task. Experimental results demonstrate that database knowledge enhances metrics and leads to more reasonable prescriptions.

**Figure 3:**
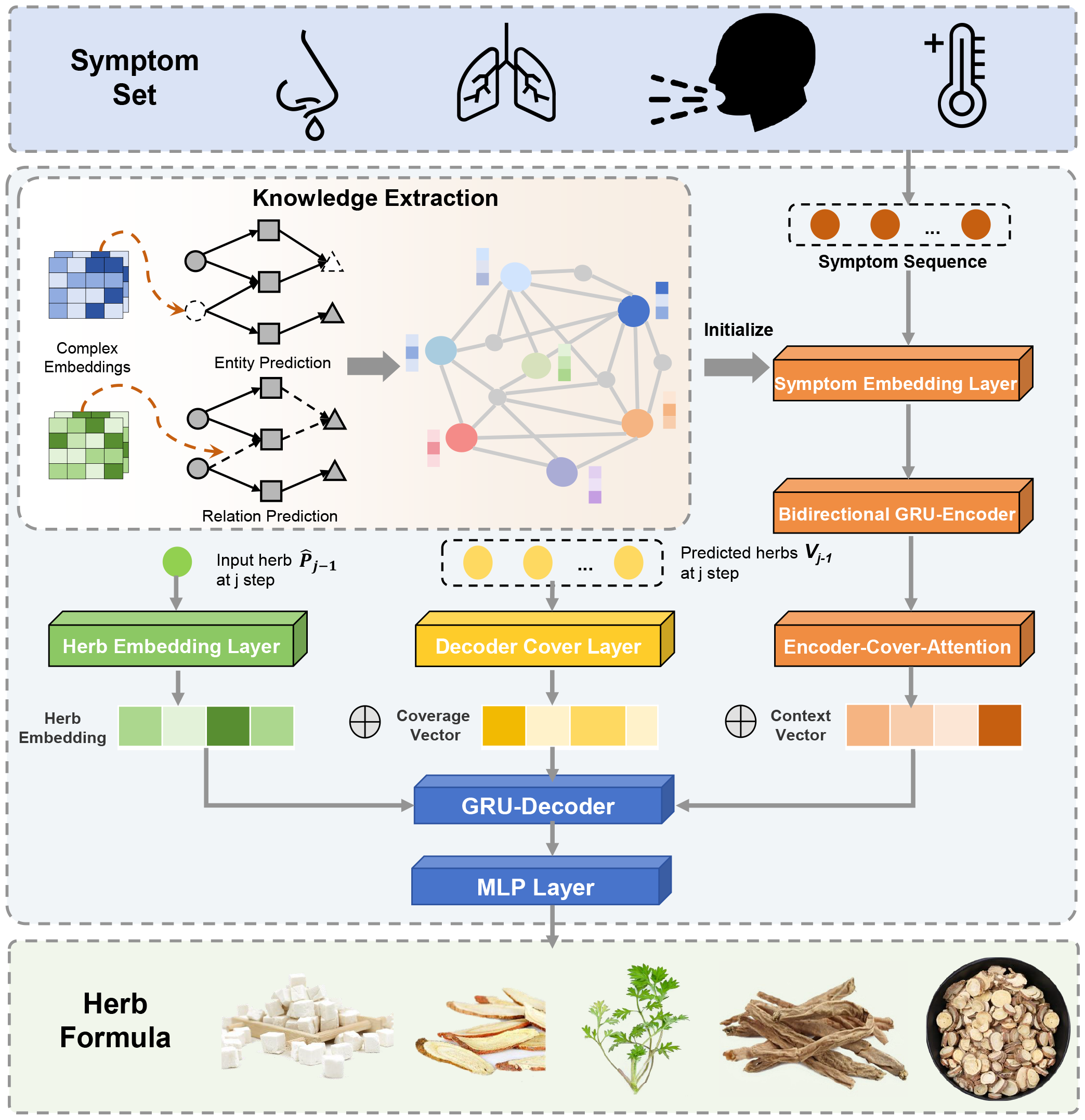
**TCM Prescription Generation Model. Knowledge Extraction** employs the ComplEX[41] model to treat ‘knowledge graph completion’ as a pre-training task, facilitating the integration of TCMM knowledge. The model first initializes the entities and relations in the knowledge graph to the complex space to acquire complex embeddings. Then, it conducts collaborative model training on two tasks: entity completion and relationship completion. The pre-trained entity embedding is then used as the initial parameter of symptom embedding layer. **Prescription Generation** model adopts a seq2seq architecture, mapping the input symptom set into a herb formula. Bidirectional GRU is utilized as the Encoder to map the pre-trained symptom embeddings into a context vector. The Decoder then progressively decodes the context vector into the herb ID *P*_*j*_. In order to prevent the generation of duplicate herbs, a multi-hot vector *V*_*j*_ is introduced to annotate the predicted herbs.

**Figure 4:**
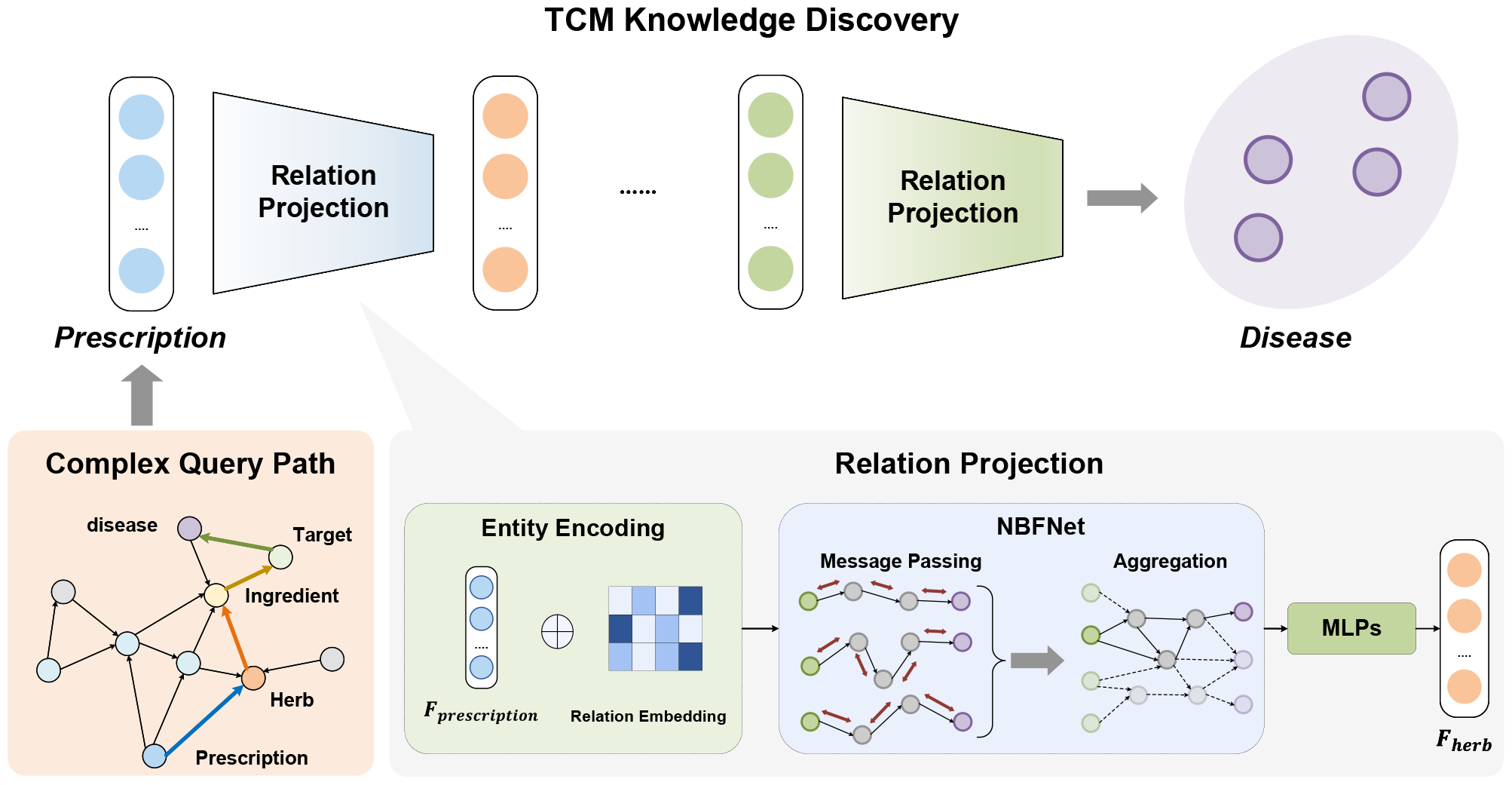
**TCM Knowledge Discovery Model** is employed to represent relations between entities within predefined complex query paths. Here, take prescription repositioning as an example, which utilizes the **complex query path** *prescription* ⟶ *herb* ⟶ *in****g****redient* ⟶ *tar****g****et* ⟶ *disease*. **TCM Knowledge Discovery** takes the fuzzy set of the source entity *F*_*prescription*_ as input and iteratively performs relation projection operations to obtain the answer set *A*_*disease*_. **Relation Projection** initially merges the fuzzy set of the head entity in the corresponding triplet with the relation embedding, then utilizes the merged information as input into the NBFNet for message passing and aggregation. Finally, the representation is mapped to the tail entity’s fuzzy set through MLP layers and sigmoid.

#### 2.3.1. Data processing

Prescription data containing dosage information is selected as the dataset to facilitate the construction of a sequence generation model from symptom sequence to herb sequence. To unify the order of sequence data, symptom sequences are sorted according to symptom ID in the database. Meanwhile, according to the principle of Jun-Chen-Zuo-Shi compatibility, the medicinal materials for treating the main symptoms usually have a larger dosage, so the herb sequences are sorted according to the weight proportion of the herbs in the prescription. The dataset contains a total of 20,911 prescriptions, which are randomly divided into training, validation, and test sets at a ratio of 8:1:1.

To clarify the relation between herbs and symptoms, the model’s initial parameters employ embeddings pre-trained on a KG. Expert knowledge directs the extraction of prescription generation-related information from the TCMM database, constructing a KG encompassing entities such as herb, prescription, ingredient, target, TCM symptom and MM symptom. The KG structure is depicted in supplementary figure S1.

#### 2.3.2. Model description

To capture the long-term dependency in symptom sequences, the GRU[44] is utilized as the Encoder to transform variable-length symptom sequences (*s*_1_, …, *s*_*M*_) into hidden state sequences (*h*_1_, …, *h*_*M*_). Additionally, the bidirectional GRU is employed to adapt to the weak order characteristics of symptom sequences, capturing symptom information before and after the current moment. The hidden state *h*_*t*_ ∈ *R*^2\**hs*^ at time t consists of the forward state *h*_*f*_ ∈ *R*^*hs*^ and the reverse state *h*_*r*_ ∈ *R*^*hs*^, with the calculation process as following:

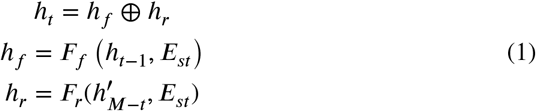

In equation 1, *E*_*st*_ represents the embedding of *s*_*t*_, while *F*_*f*_ and *F*_*r*_ are the forward GRU and reverse GRU networks, respectively 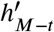 is the reverse symptom sequence.

The attention mechanism is utilized to enable the decoder to better model long sequence information of symptoms. Specifically, based on the previous state *s*_*j*−1_ and attention *A*_*j*−1_ of the symptom, the weight of the symptom hidden state at the current moment α_tj_ is measured, yielding the context vector *c*_*j*_ for the current moment of the decoder. The function a is a soft alignment function used to compute the correlation between s and h.

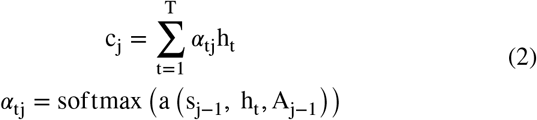

The Decoder utilizes the GRU to decode the hidden state of symptoms into a variable-length herbal sequence incrementally. Since the prescription generation task aims to generate a non-repetitive herbal sequence, a Cover layer is introduced to make the model aware of generated tokens, thereby generating a more diverse and reasonable herbal formula. The predicted herbs are transformed into a multi-hot vector representation *V*_*j*−1_, which is subsequently projected onto the hidden space *CV*_*j*−1_ via the Cover layer. This, in conjunction with the context vector *c*_*j*_, the previous state *s*_*j*−1_, and the herb embedding in the previous timestep *E*_*h*_*j*, is collectively used to update the current state *s*_*j*_ of the Decoder.

The MLP then decodes *s*_*j*_ into the herb ID *p*_*j*_.

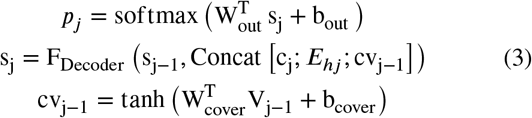

Pre-trained embedding is introduced as the initial parameter for our model to enhance the performance of downstream tasks. In order to capture the semantics within the complex graphical structure, we design a KGC task, which aims to explore high-order correlation and encode the intricate information in heterogeneous graph. ComplEX [41] is utilized as pre-trained model since its outstanding performance in the KGC task. According to [41], in order to more fully integrate the knowledge within the graph, both entity prediction and relation prediction are jointly used as training tasks.

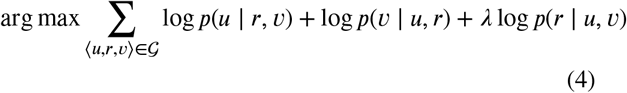

As the equation 4 shown, the objective of ComplEX is to maximize the predictive probability of the combined optimization equation, encompassing three tasks: predicting the head entity u, the tail entity v, and the relation r.

Cross entropy is used as the loss function of the model. However, the original cross entropy function requires that the target sequence have a strict order in order to measure the loss of the predicted sequence. Since the herb sequence in the prescription generation task does not demand a strict order, this work employs the smoothed target probability 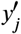 to relax the penalty for incorrect prediction of herb position.

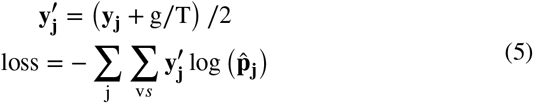

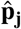 is the predicted probability that the herb appears at the jth position of the prescription. *y*_*j*_ is the one-hot encoding of the herb at the jth position of the real prescription sequence. g is the multi-hot encoding of the herbs in the real prescription; if the herb appears in the real prescription, the element at the corresponding position of the g is 1, otherwise it is 0. T is the length of the real prescription and vs represents the vocabulary size.

To support the online inference, it is essential to ensure that the result is globally optimal and has a lower time complexity. Diverse Beam Search[43] strikes a balance between greedy search and exhaustive traversal, dividing the search space into G Beam Search groups, retaining the B sequences with the highest probability as candidates in each iteration, and introducing diversity penalty to enhance sequence diversity.

#### 2.3.3. Experimental study

To validate the enhancement resulting from the incorporation of different types of modernized TCM knowledge into the prescription generation model, we conduct two comparative experiments.

The “Basic” model initializes embeddings arbitrarily without incorporating additional knowledge. The CPMCP database, which has the richest variety of relationship types among TCM databases, is utilized for comparison. The ‘CPMCP-based’ model is pre-trained using relation information from CPMCP. The ‘TCMM-based’ model is pretrained with a KG (Supplementary Figure S1) extracted from TCMM, which contains a more diverse types of relations compared to CPMCP.

Regarding hyperparameters, the maximum epoch is set to 100 and an early stopping strategy is employed. The batch size is set to 128, the embedding dimension is 600, and the hidden dimension is set to 300. The Adam optimizer is chosen for optimization, with a learning rate of 0.001. Precision, Recall, and F1 score are utilized as evaluation metrics, supplementary table S2 shows the results.

The experimental results show that the introduction of additional TCM knowledge will significantly improve the model effect. Moreover, the model based on TCMM knowledge is better than the ‘CPMCP-based’ model in terms of Precision and F1 score for prescription generation, which means that a more comprehensive modernized TCM knowledge can help identify more symptomatic herbs, thus validating the potential of the TCMM database.

The evaluation metrics are based on a statistical analysis of actual prescriptions extracted from the test dataset.

### 2.4. Multi-hop reasoning based TCM knowledge discovery

Knowledge graph reasoning can infer new knowledge or query answers based on existing knowledge and has been widely applied in biomedical tasks such as drug-target interaction(DTI) and drug-drug interaction(DDI). However, the lack of direct relational data in the current TCM field makes it challenging for traditional knowledge graph reasoning methods to solve complex queries. Multihop reasoning, as a knowledge graph reasoning task, is primarily utilized to answer complex first-order logic (FOL) queries involving logical operations such as existential quantifier (∃), conjunction (∧), disjunction (∨), and negation (¬). Therefore, in this work, we introduce a multi-hop reasoning based pipeline in Figure 8 to transform knowledge discovery into complex logical queries, discovering new knowledge through specific meta-paths.

#### 2.4.1. Data processing

To facilitate comparison with biological experiments, this work primarily concentrates on two tasks: prescription-disease and symptom-target. We extract relations from the database and construct a KG with task-relevant information specified based on expert knowledge. In contrast to the prescription generation task, which focuses on TCM-related attributes such as tropism and flavour, we retain the attribute entities of Western medicine, including biological process and pharmacological class, in order to investigate the correlation between TCM entities and Western medicine entities. Detailed information can be found in supplementary figure S2. We generate training data based on the method stated in [45], and introduce certain constraints, that filter out multiple self-related loops between entities and prohibit the appearance of bidirectional relations in the same path, to ensure the rationality of the path. In addition, logical operations, such as disjunction (∨), and negation (¬), are not considered based on the path features of downstream tasks. Furthermore, since the downstream tasks involve 4p and 5p queries, 4p and 5p data is utilized for training to enhance performance. In order to balance the performance of each query, the quantity of 4p and 5p is limited to one-tenth for other queries because the amount of answers for these tasks is significantly higher than others. The dataset is shown in supplementary table S3.

#### 2.4.2. Model description

GNN-QE[46], as the current state-of-the-art in the complex logical query task, is chosen to complete this task. This model predicts the fuzzy set of answer entities given the head entity and relation, where the elements in the set are all entities on the KG and are represented by a probability for the confidence of the tail entity.

GNN-QE initially transforms an FOL query into an expression of fundamental operations, enabling answer retrieval via expression execution. For example, FOL queries are decomposed into the expression6.

*P*_*r*_(*x*) represents the tail entity fuzzy set measured with the relation r and the fuzzy set x of the head entity. {*prescription*} represents the entity sets related to the prescriptions.

Complex logical query on the KG attempts to predict the fuzzy set of the tail entity given the fuzzy set of the head entity x and the relation r. However, traditional KGC methods based on GNN concentrate on the relation projection between single entities, which is difficult to extend to large-scale fuzzy set prediction of the high time complexity. Therefore, NBFNet[47] is utilized to model relation projection in this study. Based on the generalized Bellman-Ford algorithm for single-source problems on graphs, NBFNet has been shown to perform well in the GNN-QE framework due to its lower complexity.

Following the NBFNet, we utilize the function to incorporate the probabilities in the head entity fuzzy set *h*_*v*_ into the relation embedding, which together serves as the entity’s initial representation. The entity representation is then input into a multi-layer GNN, integrating the structural information in the graph through message passing and message aggregation. The output of the final layer is then passed to the MLP layer, which predicts the fuzzy set of the tail entity using the sigmoid function. Based on the multi-hop relations in the path, multiple iterations are performed

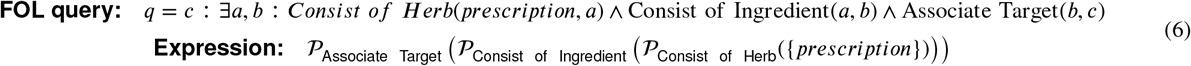

to ultimately predict the fuzzy set of answers. In equation 7, F represents the function that integrates the tail entity fuzzy set *P*_*r*_(*x*) of the previous hop and relation embedding. *ℰ*(*v*) is the triplets of the training KG.

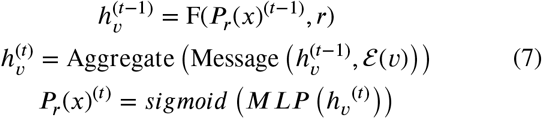

According to [47], we choose binary cross entropy loss to train our model. Ans represents the answer set of the multi-hop query, *V* is the set of all entities and *y*_*i*_ represents the probability of i in the final answer fuzzy set.

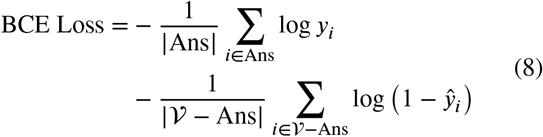

#### 2.4.3. Experimental study

Prior work[48, 49, 45, 46] of multi-hop reasoning mainly concentrated on evaluating the performance of 1p,2p,3p queries. However, this study focuses on long-path reasoning. To verify the impact of introducing 4p and 5p data to the model, we conduct an ablation study. ***GNN*−*QE***_***org***_ refers to the basic model, with the dataset presented in supplementary table S3 and ***GNN* − *QE***_***short***_ removes the 4p and 5p data from trainset. To compare model performance, both models employ the same hyperparameters, which are shown in supplementary table S4.

The performance is measured by mean reciprocal rank (MRR). According to the results present in supplementary table S5, the use of long path data demonstrates an enhancement in the performance for queries involving 3p, 4p, and 5p, while it will decrease the performance on short path queries. However, since TCM knowledge discovery requires the integration of intermediate information, it is necessary to have high performance in long-path reasoning.

## 3. Results

### 3.1. Modernized TCM research via web interface

TCMM presents a user-friendly website shown in Figure 5, enabling users to effortlessly access comprehensive information and relations among various entities in both TCM and modern medicine. To enhance user experience, the website focuses on showcasing the nine most frequently used entities: prescription, medicinal material, ingredient, pathway, gene, disease, TCM symptom, MM symptom, and syndrome.

**Figure 5:**
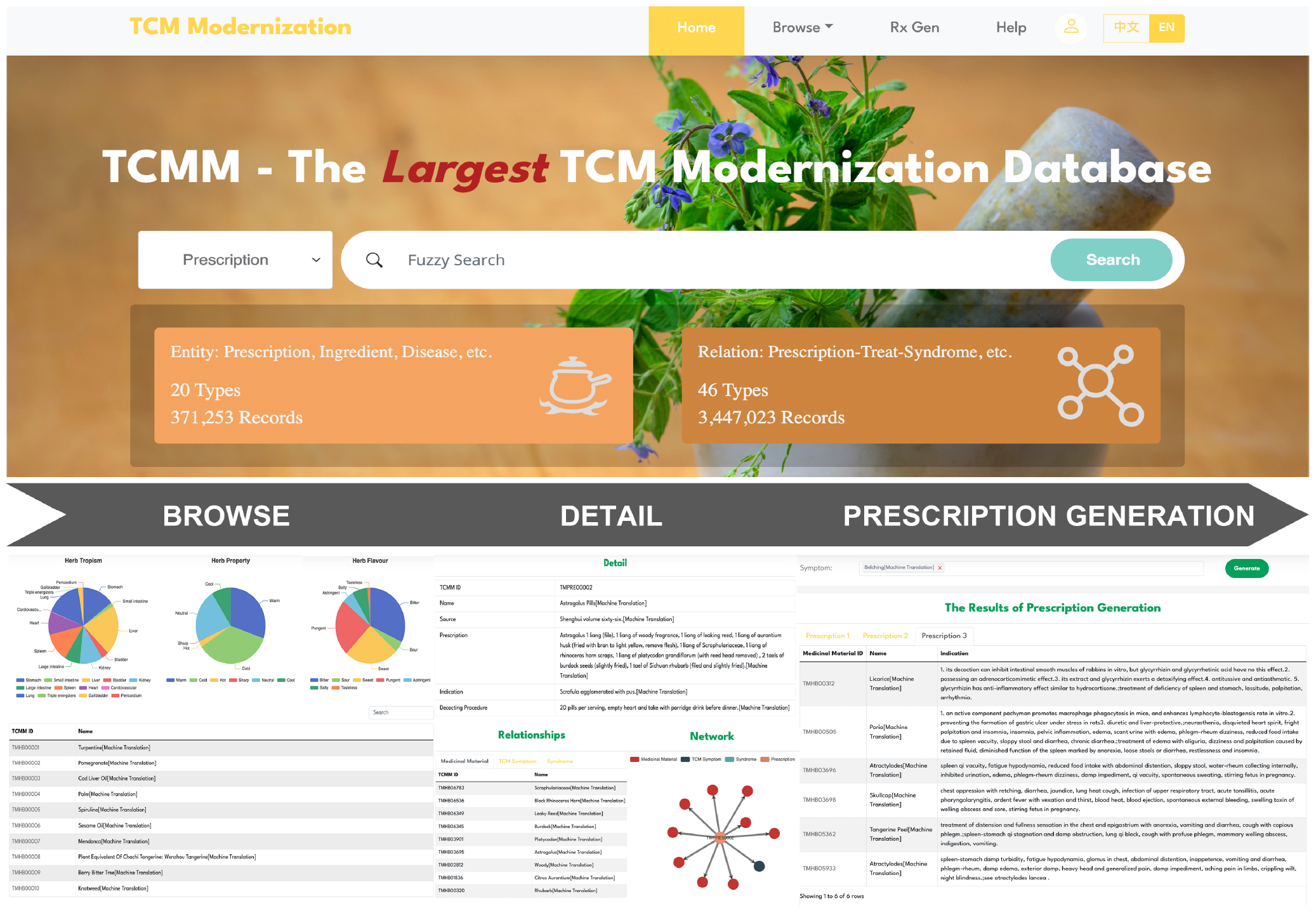
**Overview of Web Interface. Home Page** contains a navigation bar and a multi-entity search function. **Browse** shows the detailed information and statistical results of each type of entity. **Detail** displays the entity’s attributes and the associated relation types in the format of network and table. **Rx Gen** integrates the function of customized prescription generation. Users can get the top-3 prescriptions by entering the combination of the predefined 1402 symptoms.

The homepage of TCMM is equipped with a multi-entity search function, a concise description of the database, and a navigation bar filled with diverse functions. The search function is designed to support fuzzy search, allowing users to perform searches in Chinese, Pinyin, English, or using Alias names.

The Browse section is a collection of detailed information and statistical results pertaining to the entities. Users can click on the ID within the details to view the relations of the entity. The Relation section is split into three parts: detail, network, and relationships. The Detail primarily shows the entity’s attributes, while the network and relation part displays the relation types associated with the entity in the form of a knowledge graph and a table, respectively.

A key function of TCMM is Rx Gen, which is used for online prescription generation. Users can freely combine any of the predefined 1402 symptoms to perform online reasoning. The website is programmed to automatically filter and display the top three prescriptions, each containing detailed information about the medicinal material, thereby guiding the diagnosis and treatment in TCM.

### 3.2. Case studies

#### 3.2.1. TCM prescription generation

Beyond comparing model performance, it is necessary to validate the feasibility of generating prescriptions based on different types of modernized TCM knowledge. To validate the efficacy of the generated prescriptions, we randomly select various samples from the test set and compare the prediction results of the three models with ground truth.

In Table 2, herbs that appear in actual prescriptions are highlighted in green, herbs that do not appear in actual prescriptions but are symptomatic are highlighted in blue, and herbs not related to symptom are highlighted in red. Following the design of the model performance comparison experiment, ‘Basic’ represents a model without any additional knowledge incorporated. ‘CPMCP-based’ signifies a model that includes relation information from CPMCP database. The ‘TCMM-based’ model integrates relevant knowledge extracted from TCMM. According to Table 2, the pre-train models have a greater tendency to predict symptomatic herbs than the model with random initialization, even if these herbs are absent from actual prescriptions. Meanwhile, the model trained with the TCMM KG is able to identify a greater amount of symptomatic herbs and fewer irrelevant herbs than the model trained with the CPMCP relations. This further validates the effectiveness of the abundant entity types and detailed relation types present in TCMM for prescription generation task.

**Table 2.**
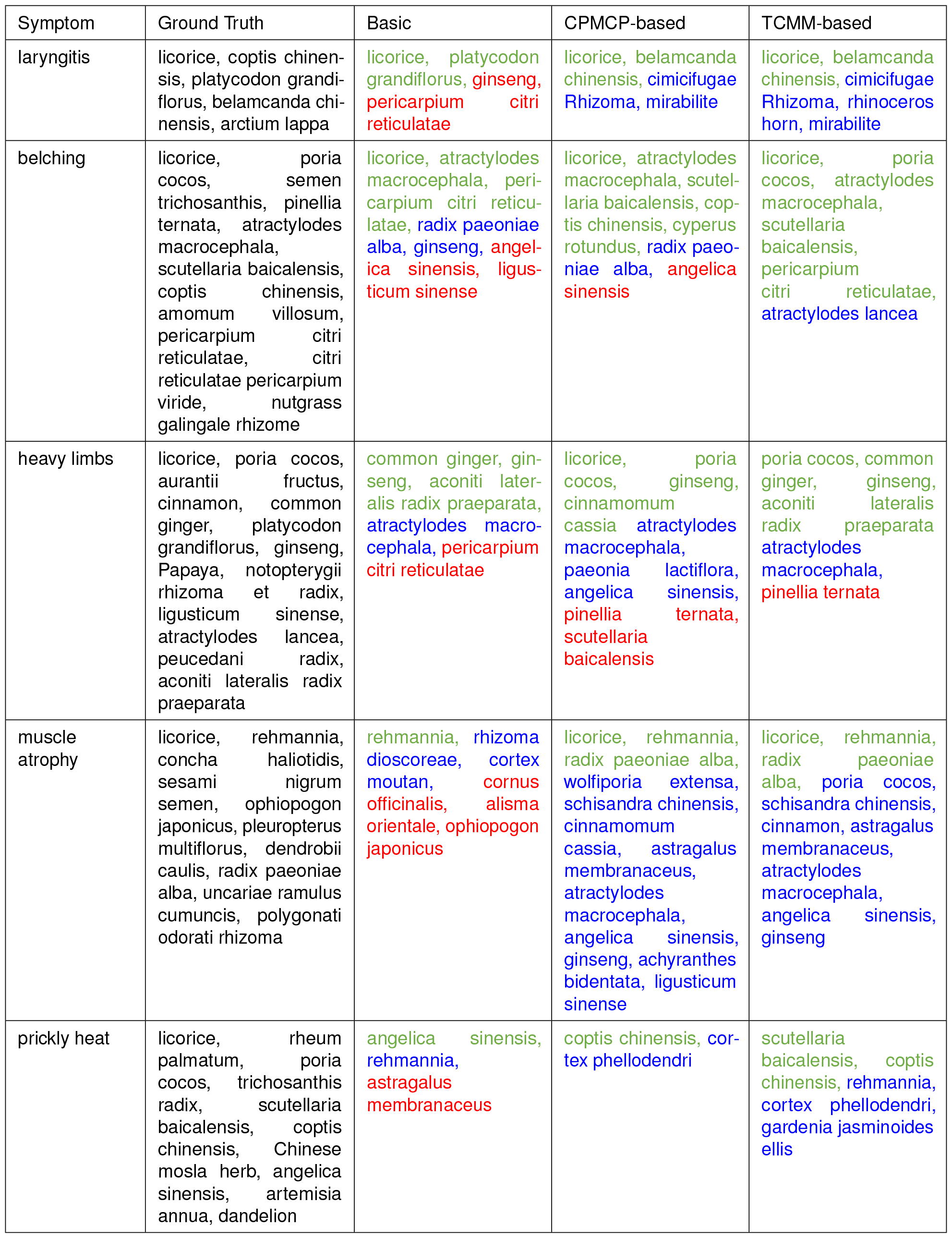
**Case Study of Prescription Generation** is used to validate the efficacy of the generated prescriptions. Herbs used in actual prescriptions are marked in green, while herbs not in the prescriptions but symptomatically relevant are in blue, and those unrelated to symptoms are in red. Besides, ‘Basic’ represents a model without any additional knowledge incorporated. ‘CPMCP-based’ signifies a model that includes relation information from CPMCP database. The ‘TCMM-based’ model integrates relevant knowledge extracted from TCMM.

#### 3.2.2. TCM knowledge discovery

We evaluate the effectiveness of multi-hop reasoning in knowledge discovery for TCM modernization with a series of cases. To discover new insights, we only consider entities outside the training KG as the answer set. Meanwhile, for comparing the model outputs with actual experimental results, we choose prescription repositioning and symptom-related target prediction as downstream tasks, which have extensive experimental cases.

Table 3 shows the case study of prescription repositioning. In order to guarantee the validity of the path data, we choose 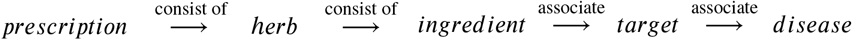 as the inference path based on the expert knowledge. We focus on five prescriptions with primarily concentrated components and select the top five answer entities based on their scores for analysis. The results demonstrate that the model’s predictions of high-scoring outputs are highly consistent with the biological experimental results. Additionally, some modern medical insights distinct from TCM knowledge have been discovered. For instance, Salvia miltiorrhiza is primarily used in TCM to treat heart disease, but its active ingredient, Tanshinone IIA, has recently been shown to be effective in cancer treatment[50]. Another example is Panax notoginseng, whose main function is to promote blood circulation, and is frequently used to treat traumatic injuries. Its ingredient notoginsenoside R1 has been proven to be effective against pressure overload-induced cardiac hypertrophy, and the model associates it with aortic stenosis, the related symptom of pressure overload-induced cardiac hypertrophy. Therefore, TCMM can be utilized to predict the potential biological activities of natural products in various herbs, contributing to the development of natural medicinal chemistry and accelerating the discovery of investigational new drugs.

**Table 3.** **Case Study of Prescription Repositioning** shows the matching degree between the model’s inference results and the experimental results of Prescription Repositioning task. If an item is marked in red, it indicates the consistency with the experimental results. ‘Publication’ shows the source of the experimental results.

| Prescription | Disease | Publication |
| --- | --- | --- |
| **Galculus Bovis Capsules** | sarcomatoid mesothelioma 0.31<br>ampulla of vater adenocarcinoma 0.31<br>stomach carcinoma in situ 0.3<br>ecthyma 0.3<br>linitis plastica 0.3 | [51] |
| **Danshen Granules** | salivary gland neoplasm 0.42<br>intestinal neoplasm 0.41<br>lung adenocarcinoma 0.41<br>neurotic depression 0.41<br>breast fibrocystic disease 0.4 | [50] |
| **Patrinia Soup** | cutaneous adenocystic carcinoma 0.3<br>splenic neoplasm 0.3<br>lung adenoid cystic carcinoma 0.29<br>acanthoma 0.29<br>pleomorphic lipoma 0.29 | [52] |
| **Panax Notoginseng Cream** | diarrheal disease 0.35<br>aortic stenosis 0.32<br>spindle cell neoplasm 0.32<br>sebaceous gland disease 0.31<br>dental abscess 0.31<br>dentin dysplasia type II 0.3 | [53] |
| **Chuanbei Powder** | functional gastric disease 0.35<br>dentin dysplasia type II 0.3<br>fish eye disease 0.3<br>tarsal tunnel syndrome 0.29<br>deficiency anemia 0.28 | [54] |

Inflammatory response as a common symptom has been extensively studied, and the understanding of its gene regulation mechanism significantly benefits modern medical diagnosis and treatment. Therefore, for the case study of symptom-related target prediction, we adopt 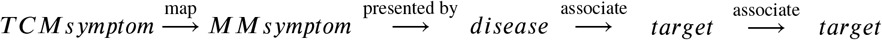 as the reasoning path and mainly analyze symptoms related to inflammation. The results in

Table 4 show that the model can effectively predict symptom-related regulatory genes. For example, GABARAPL2, as a specific protein, is preferentially recognized by autoantibodies from early rheumatoid arthritis patients[55]. This kind of prediction may provide valuable information for TCM researchers to identify the underlying molecular basis of patient symptoms.

**Table 4.** **Case Study of Symptom Related Target Prediction** shows the matching degree between the model’s inference results and the experimental results of Symptom Related Target Prediction task. If an item is marked in red, it indicates the consistency with the experimental results. ‘Publication’ shows the source of the experimental results.

| Symptom | Target | Publication |
| --- | --- | --- |
| **Rheumatoid Arthritis** | GABARAPL2 0.34<br>IQCJ-SCHIP1 0.34<br>TMEM33 0.33<br>SORT1 0.33<br>SH3GLB1 0.33 | [55] |
| **Pneumonia** | USP50 0.29<br>STX12 0.29<br>NDUFA6 0.29<br>APEX2 0.28<br>VAMP4 0.28 | [56] |
| **Tracheitis** | CREB3 0.4<br>PSMC5 0.4<br>TFAP2C 0.4<br>FRMD1 0.39<br>SNTB2 0.39 | [57] |

## 4. Discussion

Based on information regarding TCM prescription, symptom, target, and ingredient, numerous TCM databases have been developed using various data sources. Unfortunately, current efforts to clarify the pharmacological mechanisms of TCM are hindered by the absence of correspondence between TCM and Western medical knowledge. To address these issues, we conduct a study that integrates existing TCM and Western medical databases, refining the relationships between the two fields through a standardized approach.

In order to enhance data quality, a rigorous screening and verification process is implemented for database entities. Specifically, we employ a data alignment process that filters entities based on specific rules, such as ID, name, description, and other relevant information. We also merge duplicate entities within the existing database to enrich entity information. For example, aliases are used to consolidate identical medicinal materials, and LLM is applied to merge semantically similar symptoms. These measures ensure that the database encompasses the majority of entities and relations in TCM and Western medicine, greatly enhancing its practicality in discovering modernized TCM knowledge. Moreover, the potential of the TCMM is validated from two perspectives. By integrating modernized TCM knowledge, the TCM prescription generation method is strengthened, resulting in prescriptions that are not only highly consistent with classical TCM prescriptions but also include new effective components to enhance the therapeutic effects of the original prescriptions. The field of TCM knowledge discovery is limited by the lack of relational data, leaving a gap in the use of computational methods to mine TCM knowledge. Existing methods typically rely on network pharmacology, with a singular and limited focus on specific prescriptions and diseases. The introduction of the TCMM database effectively bridges the gap between TCM and Western medical research, making AI-based, generalized TCM knowledge discovery possible. This study selects prescription repositioning and symptom-related target identification tasks with abundant cases for result validation. Experiments demonstrate that novel pathways consistent with the results of biological experiments can be discovered using TCMM knowledge, further proving the immense potential of TCMM in TCM knowledge discovery.

However, despite our experiments demonstrating the potential of TCMM knowledge, there are still some limitations. First, the database requires further improvements, particularly in the information of prescription. TCMM employs a hybrid approach that combines rules and LLM to extract relations among prescription, medicinal material, and symptom from text. Although the model achieves high accuracy in parsing prescriptions, extensive manual verification is still necessary to standardize the results, which will be addressed in future updates. Moreover, while TCMM data has bridged the knowlegde between TCM and modern medicine, enabling the discovery of modernized TCM knowledge. However, long-path logical queries introduce a large number of random variables, unavoidably reduces the accuracy of the results. Additionally, the training data has not been completely validated by biological experiments, containing noise. Therefore, future research will continue to integrate the latest studies, improve the authenticity of the data, such as target and disease-related information, supplementing graph data to shorten inference paths and enhance the confidence of new knowledge. TCMM will be regularly updated to ensure its ongoing relevance and performance, accelerating the progress of modernized TCM research.

## 5. Conclusion

In conclusion, the TCMM database represents a ground-breaking advancement in TCM research, offering the largest collection of modernized TCM data and deep learning integration. This resource enhances our understanding of TCM, promotes drug development, and aids clinical applications, setting a new standard for the fusion of traditional wisdom and modern science.

### CRediT authorship contribution statement

**Zhixiang Ren:** Conceptualization, Formal analysis, Supervision, Funding acquisition, Writing review editing. **Yiming Ren:** Methodology, Model training, Writing-original draft. **Zeting Li:** Methodology, Data curation, Model training. **Huan Xu:** Conceptualization, Supervision, Funding acquisition, Writing-review editing.

### Declaration of competing interest

The authors declare that they have no known competing financial interests or personal relationships that could have appeared to influence the work reported in this paper.

## Supporting information

Supplemental materials, and will be used to illustrate the results of the manuscript

## Data availability

Main data and related functions are publicly available through a web interface at https://www.tcmm.net.cn/. To obtain the complete data, please contact the corresponding author.

## Funding

This work is funded by National Natural Science Foundation of China [grant No. 42177417]. This work is also funded by Shanghai Frontiers Science Center of Optogenetic Techniques for Cell Metabolism (Shanghai Municipal Education Commission).

## Acknowledgements

The study is supported by the Peng Cheng Laboratory and Peng Cheng Cloud-Brain.

## Supplementary Materials

Supplementary material to this article can be found online.

## Notes

### Competing Interest Statement

The authors have declared no competing interest.

### Funding Statement

This work is funded by Shanghai Frontiers Science Center of Optogenetic Techniques for Cell Metabolism (Shanghai Municipal Education Commission).

