## Supplemental materials, and will be used to illustrate the results of the manuscript for "TCMM: A Unified Database for Traditional Chinese Medicine Modernization and Therapeutic Innovations"

Table of Contents

| **Table S1** | S2 |
| --- | --- |
| **Table S2** | S2 |
| **Table S3** | S2 |
| **Table S4** | S3 |
| **Table S5** | S3 |
| **Figure S1** | S4 |
| **Figure S2** | S5 |

**Table S1. Medicinal Dosage Conversion** depicts the alignment method for dosage information in prescriptions. Measurement information within prescription descriptions has been standardized to grams (g) and milliliters (ml).

| 1 liang | 1 qian | 1 zhu | 1 fen | 1 li | 1 hao | 1 jin | 1 gongjin | 1 he | 1 dou |
| --- | --- | --- | --- | --- | --- | --- | --- | --- | --- |
| 31.25g | 3.125g | 1.3g | 0.3125g | 0.03125g | 0.003125g | 500g | 1000g | 20ml | 2000ml |

**Table S2.** **Prescription Generation Result.** Test results of the prescription generation model, which benefits from better database knowledge.

|  | **Precision** | **Recall** | **F1 score** |
| --- | --- | --- | --- |
| **Basic**  **CPMCP-based**  **TCMM-based** | 26.774  26.571  **27.446** | 19.882  **20.771**  20.745 | 22.819  23.316  **23.629** |

**Table S3.** Statistics of different query types utilized in the multi-hop reasoning dataset

|  | **1p** | **2p** | **3p** | **4p** | **5p** |
| --- | --- | --- | --- | --- | --- |
| **Train**  **Test**  **Valid** | 249596  86900  86773 | 249596  7000  7000 | 249596  7000  7000 | 25000  7000  7000 | 25000  7000  7000 |

**Table S4. Hyperparameter of GNN-QE** are selected by the performance on the validation set.

| **Hyperparameter** | | **Values** |
| --- | --- | --- |
| **GNN** | Number of layers  Hidden dimensions | 4  32 |
| **MLP** | Number of layers  Hidden dimensions | 4  32 |
| **Traversal Dropout** | Probability | 0.25 |
| **Learning** | Batch size  Sample weight  Optimizer  Learning rate  Batch per epoch  Adv. temperature | 24  uniform across queries  Adam  5e – 3  300,000  0.2 |

**Table S5.** Test MRR result (%) of GNN-QE. ${GNN-QE}_{tcmm}$ is the model trained with 4p and 5p data, while ${GNN-QE}_{short}$ is not.

|  | **1p** | **2p** | **3p** | **4p** | **5p** |
| --- | --- | --- | --- | --- | --- |
| ${GNN-QE}_{tcmm}$  ${GNN-QE}_{short}$ | 13.9  **14.6** | 1.8  **1.9** | **1.8**  1.6 | **3.1**  2.9 | **3.9**  3.4 |

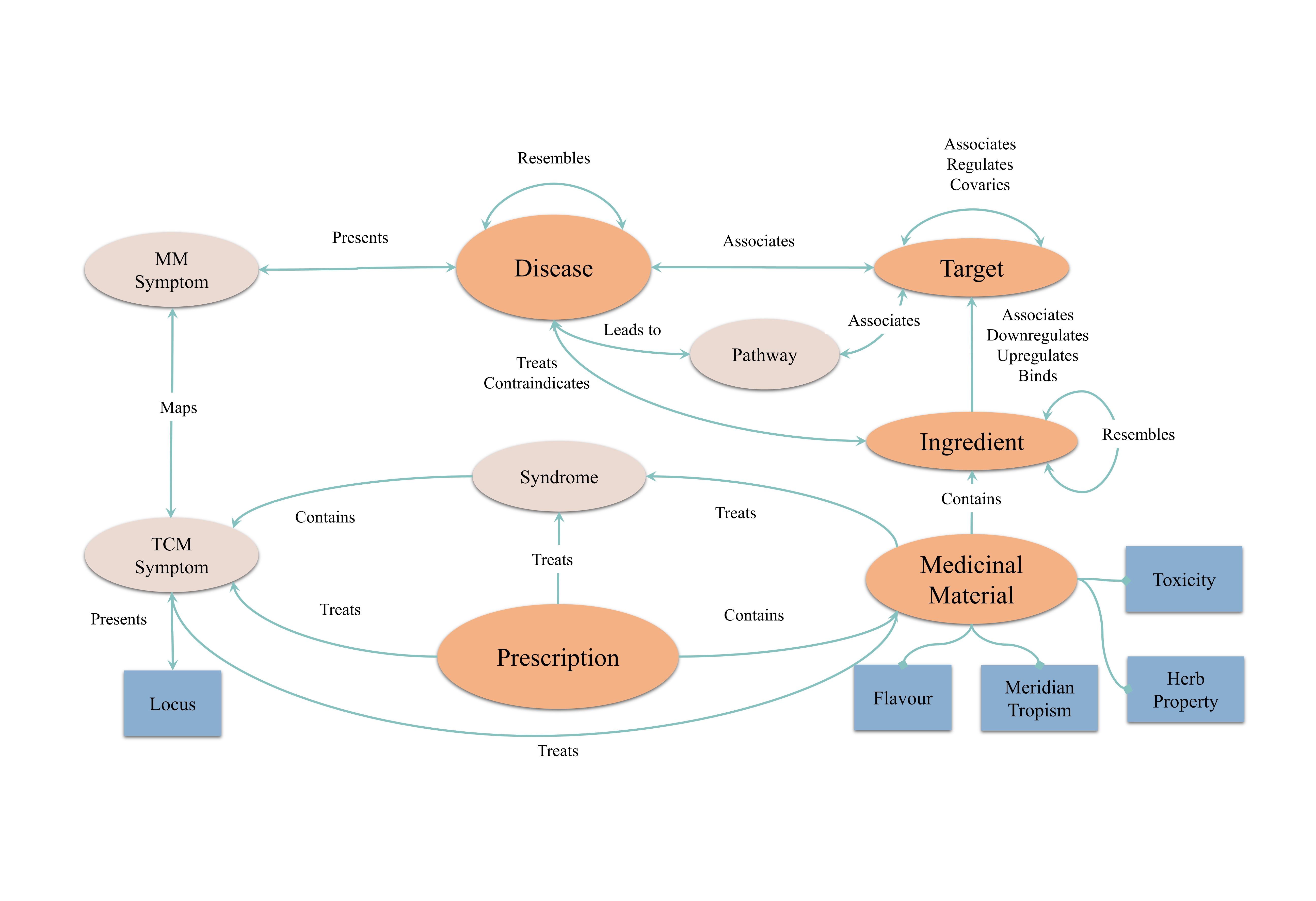
**Figure S1. Knowledge Graph for Prescription Generation** displays the knowledge graph used by the prescription generation model for knowledge extraction. The attributes of medicinal material and prescription information are retained to characterize TCM compatibility principles. In addition, modern medical information such as target, disease, ingredient, and pathway are integrated to assist this task.

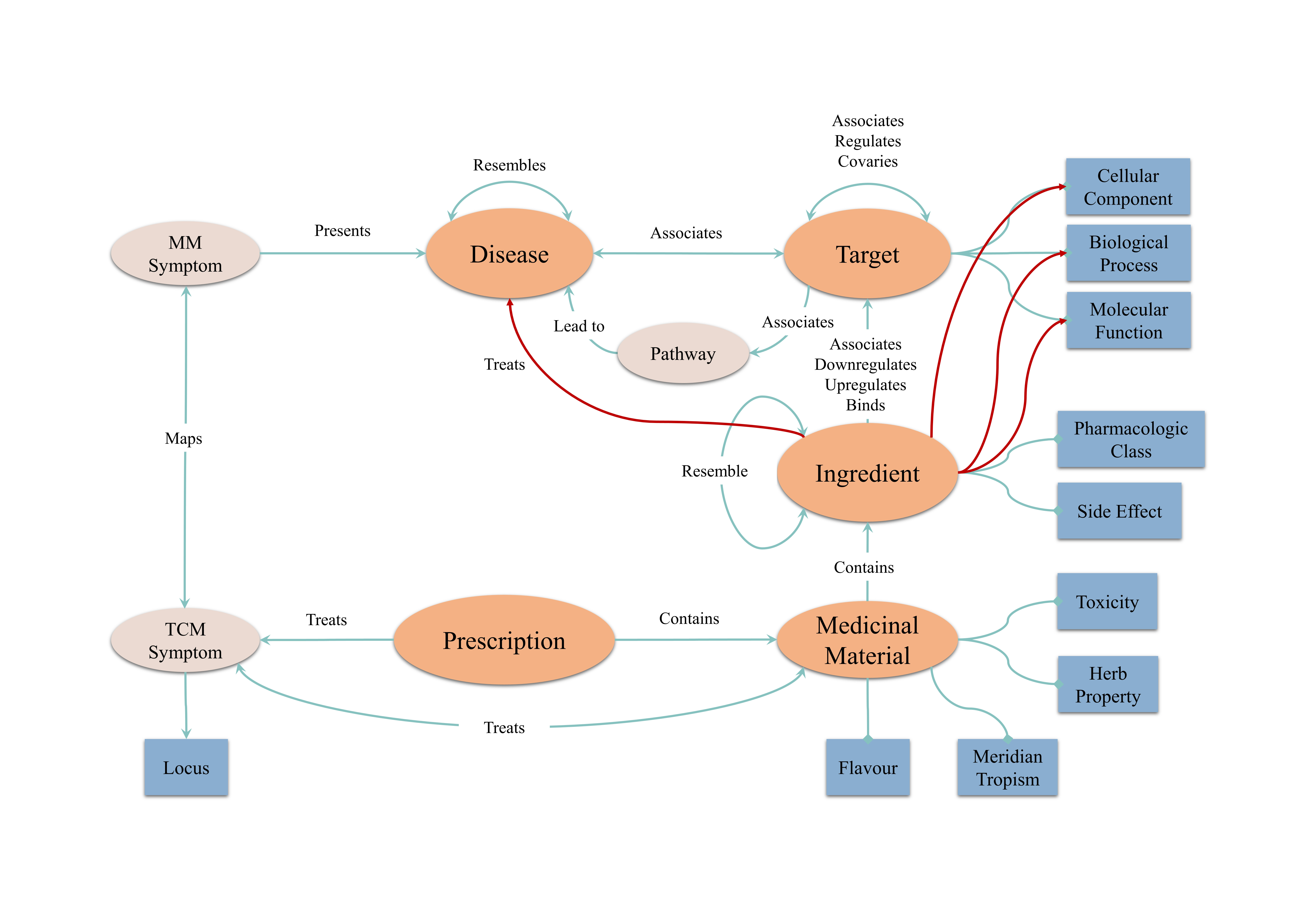
**Figure S2. Knowledge Graph for Knowledge Discovery** presents the knowledge graph utilized by the knowledge discovery model. To fully explore the correlation between TCM and Western medicine, only entities like anatomy and syndrome are removed, while all attribute information is preserved.
